# Comparative Computed Tomography with Stress Manoeuvres for Diagnosing Distal Isolated Tibiofibular Syndesmotic Injury in Acute Ankle Sprain: a Protocol for an Accuracy-Test Prospective Study

**DOI:** 10.1101/2020.01.24.20018515

**Authors:** João Carlos Rodrigues, Alexandre Leme Godoy dos Santos, Marcelo Pires Prado, José Felipe Marion Alloza, Renato do Amaral Masagão, Laercio Alberto Rosemberg, Durval do Carmo Santos Barros, Adham do Amaral e Castro, Marco Kawamura Demange, Mário Lenza, Mário Ferretti

**Author notes:** Albert Einstein Hospital, Albert Einstein Avenue, 627/701, São Paulo – SP, 05652-900, Brazil, 55 11 2151-2452 or 55 11 2151-4586. ***Disclosures:*** The corresponding author and coauthors have no relevant disclosures. ***IRB:*** The internal review board and the research ethics committee (register number 62100016.5.0000.0071) approved this study, and all participants provided written informed consent.

## Abstract

**Introduction:** Although there are several imaging options for diagnosing syndesmotic injury, a fundamental question that guides treatment remains unanswered. Syndesmotic instability is still challenging to correctly diagnose, and syndesmotic disruption and true syndesmotic instability should be differentiated. Currently, imaging tests quickly diagnose the severe syndesmotic instability but have difficulty in diagnosing mild and moderate cases. This study aims to investigate which strategy among an existing computed tomography (CT) index test and two new add-on CT index tests with stress manoeuvres can more accurately diagnose syndesmotic instability. The secondary objective is to investigate the participants’ disability outcomes by applying the Foot and Ankle Ability Measure questionnaire.

**Methods and analyses:** This diagnostic accuracy-test study will consecutively select individuals older than 18 years with a clinical diagnosis of suspected acute syndesmotic injury. Three strategies of the CT index test (one in the neutral position and two with stress) will examine the accuracy using magnetic resonance imaging as the reference standard. The external rotation and dorsiflexion of the ankle will guide the stress manoeuvres. Comparison measurements between the injured syndesmosis and the uninjured contralateral side of the same individual will investigate syndesmotic instability, evaluating the rotational and translational relationship between the fibula and tibia. Sensitivity, specificity, area under the ROC curve and likelihoods will compare the diagnostic accuracies of the strategies.

**Ethics and dissemination:** The Internal Review Board and the Research Ethics Committee approved this study (registered number 62100016.5.0000.0071). All the participants will receive a spoken description of the aim of the study, and the choice to participate will be free and voluntary. Participants’ enrolment will occur after signing the written informed consent, including the terms of confidentiality. The results will be presented at national and international conferences and published in peer-reviewed journals and social media.

**Trial registration number:** ClinicalTrials.gov (identifier NCT04095598), pre-results.

**Strengths and limitations of this study:**

- This study is the first to examine the accuracy and feasibility of CT with stress manoeuvres for diagnosing syndesmotic instability.
- The disability outcomes will be used to evaluate syndesmotic instability diagnosed by CT with stress manoeuvres as a prognostic factor.
- The limitations of this study include the use of MRI as the reference standard test, which, although not perfect, is estimated to have high accuracy compared to the gold-standard arthroscopy.^1^
- An inherent degree of imprecision related to the severity of the ankle sprain and pain exacerbation may occur when the participants themselves control stress manoeuvres; therefore, some variability is unavoidable. Researchers will monitor and investigate the influence of this source of variability on the results.

## INTRODUCTION

One specific type of sprain is high ankle sprain with ligament damage to the distal tibiofibular syndesmosis. Syndesmotic lesions are the leading cause of persistent disability or chronic pain and frequently require longer recovery times and significantly more treatments compared with low lateral ankle sprains.^2,3^ The degree of instability in syndesmotic disruption guides the decision to operate or treat conservatively.^4^ Clinical examinations, routine radiographs, stress radiographs, computed tomography in neutral position (CTNP), weight-bearing computed tomography (WB-CT), and magnetic resonance imaging (MRI) are the available diagnostic tools for determining the correct treatment. The clinical diagnosis has limited evidence, and the best current practice considers that few clinical tests have any validity in identifying syndesmotic disruption.^5^ The mortise or anteroposterior radiographic view may promptly diagnose severe syndesmotic instability (SI) by identifying the widening of the articular space on the injured side relative to the contralateral unaffected side. However, SI may be underdiagnosed when routine radiography shows a usual articular relationship, and this method cannot reliably predict syndesmotic injuries.^6^ Stress radiographs are inaccurate for evaluating such injuries in a cadaveric model.^7^ Recently, WB-CT has emerged as a new modality for the examination of syndesmosis; however, studies have shown that WB-CT is not superior to CTNP,^8^ and axial loading has no benefit for the diagnosis of instability.^9^ Although MRI can visualize and diagnose syndesmotic injury with high accuracy,^10^ MRI is an expensive exam and is not widely available. CTNP is more sensitive than radiography for detecting syndesmotic widening.^11^ Nevertheless, the traditional measurement of the anterior tibiofibular distance, obtained by CTNP, has an unsatisfactory area under the receiver operating characteristic curve (AUC) performance of 0.56 for diagnosing SI.^12^ Despite all of these available methods, SI is still challenging to correctly diagnose, and syndesmotic ligament disruption and real SI should be differentiated. Comparative ankle CT with stress manoeuvres (CTSM) is an alternative approach that could improve the diagnosis of SI. To the best of our knowledge, the application of CTSM has not been previously described.

## OBJECTIVES AND HYPOTHESIS

The main aim is to investigate which strategy among an existing CTNP index test and two new add-on CTSM index tests (CTSM with extended-knees or flexed-knees) can more accurately diagnose SI. We hypothesized that the add-on CTSM with extended-knees (CTSM-EK) or CTSM with flexed-knees (CTSM-FK) would have a more accurate capability of diagnosing SI than CTNP alone. The secondary objective is to investigate participants’ disability outcomes by applying the Foot and Ankle Ability Measure (FAAM) questionnaire. We hypothesized that participants with SI, diagnosed by CTSM, will have worse disability outcomes than participants without SI.

## METHODS

This study follows the guidelines of Standards for Reporting of Diagnostic Accuracy Studies (STARD).^13^

### Study design

A single-centre diagnostic accuracy-test prospective study.

### Study setting

The Department of Radiology, in partnership with the Department of Orthopedics, conducted this study at a tertiary hospital with the approval of the Internal Review Board and the Ethics Committee.

### Participants

- Inclusion criteria
- Adults older than 18 years;
- One episode of an ankle sprain;
- Sprain episode occurred up to 3 weeks prior;
- Positive orthopaedic evaluation for suspected unilateral syndesmotic injury, defined as the presence of at least one of the following symptoms: pain during palpation of the distal tibiofibular syndesmosis; pain during the squeeze test (manual compression of the tibia and fibula in the middle third of the leg); pain during the external rotation test; and inability to stand on the tip of the affected foot.

Exclusion criteria

- Bilateral ankle sprain;
- Previous ankle surgery;
- Ankle fractures and dislocations (except avulsion fractures in ligamentous insertions or fracture of the posterior malleolus related to syndesmotic injury);
- Congenital or acquired ankle deformities; and
- Infection, inflammatory or neuropathic ankle arthropathies.

### Participant selection and recruitment

A consecutive sample of individuals with suspected syndesmotic disruption attended at the Foot and Ankle Outpatient Clinic will be referred to the Radiology Department to perform CT and MRI. A research assistant (RA) will screen all the participants for eligibility and will interview them about their willingness to participate. The RA will clarify the aims of the research and will invite those who meet the inclusion criteria to enroll. The RA will also describe all the study details, will answer all questions about the objectives, risks, benefits, and confidentiality, and will read aloud the informed consent form. Individuals who agree to participate will sign and date the informed consent form. The RA will attach a copy of the informed consent to the medical record and will hand a second copy to the participants. Participants will provide demographic data and will complete pre-exam forms before performing image exams. If for any reason, the participant is unable to sign the informed consent form, the RA will supply a verbal description of the study and will ask them to give verbal consent in the attendance of an observer who will sign the informed consent form. The reasons for exclusion or refusal to participate will be recorded. Figure 1 shows a flowchart delineating the study procedures.

**Figure 1.**
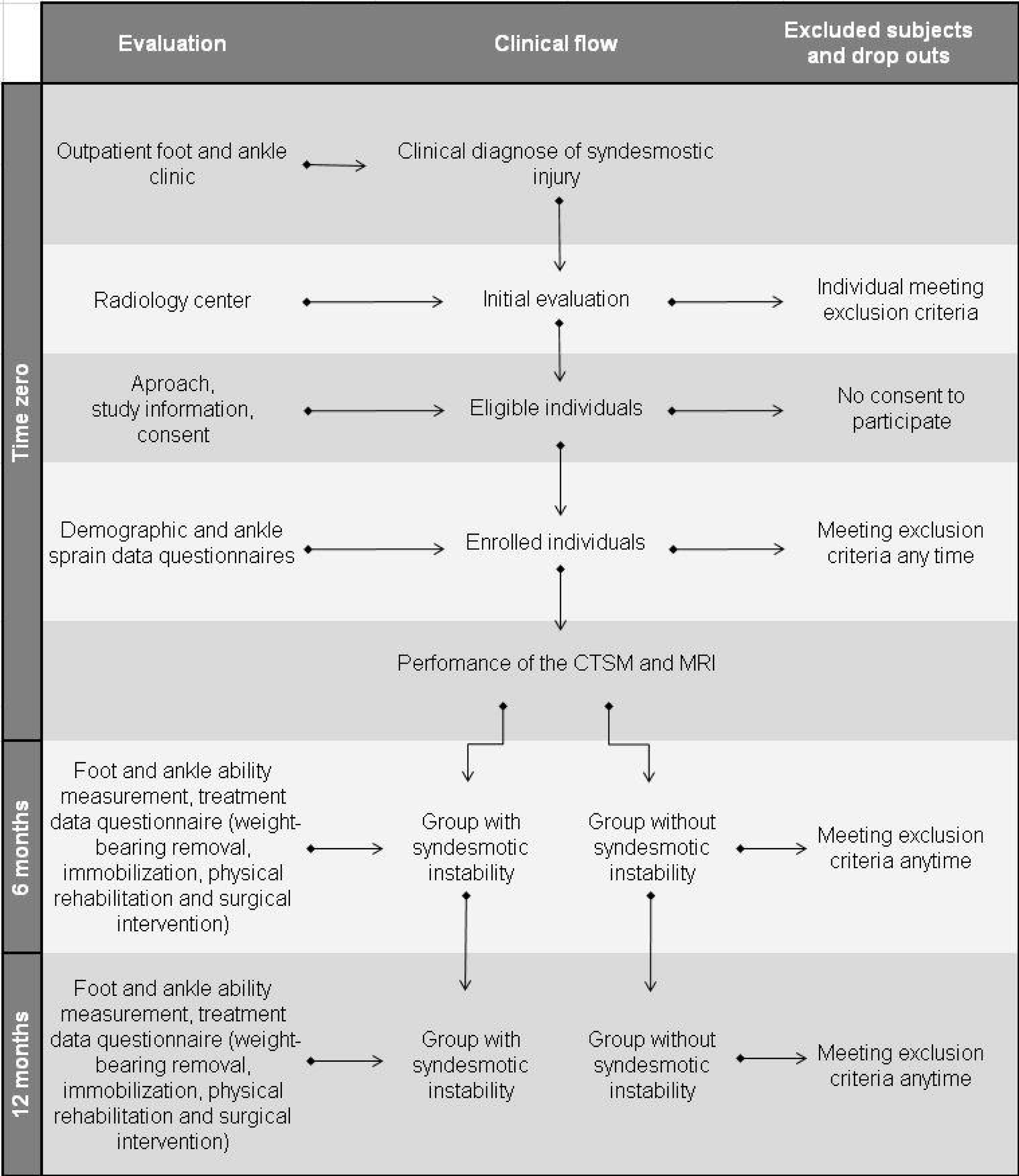
Clinical pathway for syndesmosis study.

### CT exam technical parameters

An Aquilion ONE V6 scanner (Toshiba Medical Systems, Tochigi, Japan) with 320 channels will be used to perform all the exams using the following technical parameters: a volumetric acquisition, 120 kV, 150 mA, 0.5 s rotation time, 0.5 mm slice thickness, 0.25 mm interpolation, 320-detector rows, field of view medium or large, and fine filter for bone. The lowest possible irradiation dose will produce images of diagnostic quality. The feet of the participants will be simultaneously scanned in the same field of view.

### Existing index test (CTNP)

In the neutral phase, the feet will be parallel and perpendicular to the long axis of the legs. The knees will be in an extended position.

### New index test (CTSM extended-knees)

In the first stress phase, the feet will be set in an external rotation of 45 degrees using a vertical line as a reference, and a voice command will instruct the participant to maintain maximum dorsiflexion to the limit of tolerable pain during image acquisition. The knees will be in an extended position.

### New index test (CTSM flexed-knees)

In the second stress phase, the feet will be set in an external rotation of 45 degrees using a vertical line as a reference, and a voice command will instruct the participant to maintain maximum dorsiflexion to the limit of tolerable pain during image acquisition. A support pad will keep the knees flexed at 45 degrees.

### Acrylic board

Investigators will perform the CTNP, CTSM-EK, and CTSM-FK in a standardized manner. All the participants will be scanned in the supine position with their feet supported on the acrylic board (Medintec, Mogi das Cruzes, Brazil) connected to a pair of side strings of adjustable length. The researchers will ask the participants to hold the manual support at the proximal ends of the strings and will give the participants the verbal commands through the room’s speakers to pull the strings and perform dorsiflexion at the appropriate times.

### Participants’ training stress manoeuvres and feasibility assessment

The technicians will guide participants to train dorsiflexion by simulating the movement of the feet, pulling strings just before the acquisition of the images. Difficulties in performing the stress manoeuvres, including pain exacerbation, motion artefacts, image repetition, total exam duration, and dropout will be used to assess the feasibility of the new index test.

### CTNP, CTSM-EK, and CTSM-FK reading parameters

Measurements comprising six distances, two ratios, and two angles will determine the anatomic tibiofibular relationship, examining the rotational parameters and the fibular translation concerning the incisura as described by Nault et al.^14^ Figure 2 shows the distances, ratios, and angles. A reference line 1 cm above the tibial plafond will guide the correct plane for all the measurements, except for the second angle, which will occur in the plane of the tibial plafond. Another measurement will determine the smallest distance between the fibula and the tibia in the plane of the tibial plafond, as described by Ahn et al.^12^. All the measurements will occur in a standardized way in the CTNP, CTSM-EK and CTSM-FK. Differences equal to or higher than 1 mm in distances and 2 degrees in angles between the injured and uninjured ankles of the same individual will define the cut-off for a positive test. Differences smaller than 1 mm and 2 degrees will define the cut-off for a negative test. Although there are no previous studies on CTSM, the researchers adopted cut-off points based on a recent study comparing injured and uninjured syndesmoses, which showed significant differences in the distances and angles between groups estimated at 1 mm and 2 degrees, respectively.^15^

**Figure 2.**
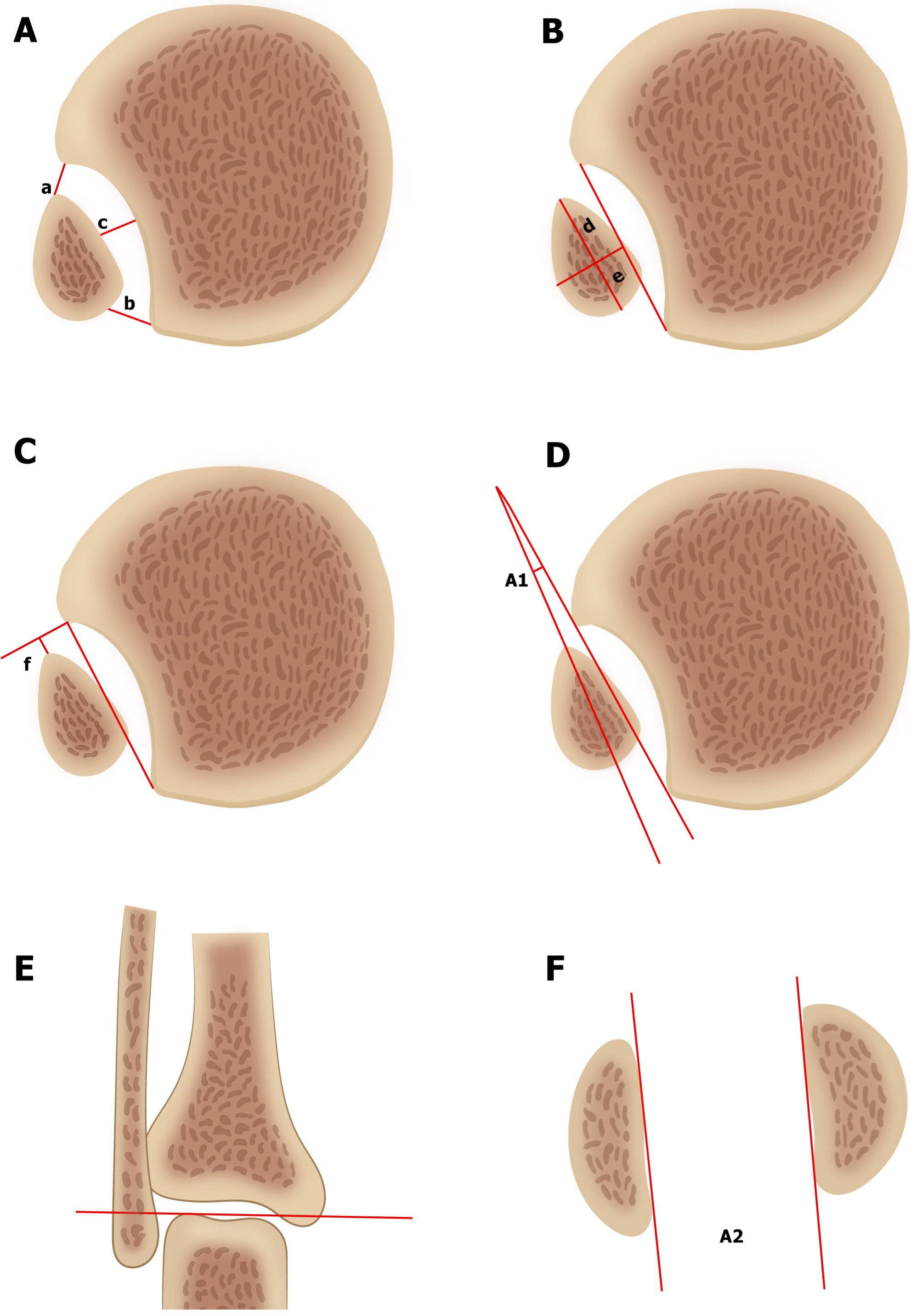
Illustrations A, B, C, and D show an axial view of a normal syndesmosis 1 cm proximal to the tibial plafond with six measures (a to f) and one angle (A1). Illustration E shows the reference line on the tibial plafond, and illustration F shows the corresponding second angle (A2) measure.

### MRI exam technical parameters

A phased array dedicated coil on 1.5-T magnet HDX (GE Healthcare, Milwaukee, USA) will be used to perform all of the examinations using the following sequences: sagittal T1-weighted (TR/TE, 542/9; number of excitations [NEX], 1; matrix, 320 × 256; thickness, 4 mm; field of view [FOV], 10 cm); sagittal T2-weighted fat-suppressed (3000/39, 2, 384 × 224, 4, 10); axial T2-weighted fat-suppressed (3483/48, 2, 384 × 224, 4, 10); coronal T2-weighted fat-suppressed (3000/39, 2, 384 × 224, 3, 10) and coronal oblique DP-weighted (2840/35, 2, 384 × 224, 3, 10).

### Reference test (MRI)

The standard protocol will acquire all of the ankle MRI for participants suspected to have syndesmotic injury as follows. Participants will be scanned with their knees in extension and ankles in a neutral position. Two studies comparing the accuracy of MRI with arthroscopy have shown that MRI is a highly sensitive and specific instrument for the evaluation of syndesmotic injury.^1,10^ The arthroscopic examination allows for correct diagnosis and treatment and is considered the best reference standard for syndesmotic injuries.^16^ However, in this study, the selection of participants is based on ankle sprain condition and physical examination, which has limited accuracy.^5^ A significant proportion of uninjured syndesmosis will be selected with the alternative diagnoses of lateral collateral ligaments injury. Applying arthroscopy to the alternative diagnoses group would have been difficult to justify ethically, considering that the index and reference tests will presumably show negative results. Even a minimally invasive procedure, such as the arthroscopic examination, may have complications and is not risk-free.^17,18^

### MRI reading parameters

The investigators prespecified the result categories, classifying syndesmotic ligaments (anterior inferior tibiofibular, posterior inferior tibiofibular, and interosseous), lateral collateral ligaments (anterior talofibular, calcaneofibular, and posterior talofibular) and deltoid ligaments (superficial and deep layer) as grade 0 (normal ligament), grade I (soft tissue oedema around the ligament but still intact), grade II (partial tear with high signal intensity and thickening) or grade III (complete ligament tear with avulsion or discontinuity) injury.^19^

### Foot and Ankle Ability Measure questionnaire

During the follow-up, the RA will contact participants six and 12 months after CT and will ask them to answer the FAAM questionnaire by phone or online. The FAAM questionnaire will quantify disability.^20^ The FAAM is a reliable, responsive, and valid tool for evaluating physical activity for a variety of disorders of the leg, foot, and ankle. A systematic review of the literature identified the FAAM as one of the most appropriate patient-assessed tools to quantify functional ankle disabilities.^21^ The FAAM questionnaire consists of two subscales with 29 questions in total. The Activities of Daily Living (ADL) subscale has 21 questions and assesses an individual’s ability to stand, squat, or climb stairs. The Sports subscale has eight questions and assesses the ability to run, jump, and land. The two subscales are reported separately, with a score range of 0-84 for the ADL subscale and 0-32 for the Sports subscale. The scale range for each subscale changes from the lowest score (zero) to the highest (100) in percentage. Higher scores represent an improvement in the outcome for both subscales. The exposure factor of interest will be SI detailed in the item CTNP, CTSM-EK, and CTSM-FK reading parameters. Data will be collected on the following covariates related to the participants’ characteristics: age (years), gender (male or female), and body mass index (BMI). Data will also be collected on the covariates related to treatment: weight-bearing restrictions (partial or total), immobilization (cast or robot-foot), physical rehabilitation (number of sessions), and surgical intervention (screws or suture-bottom fixation). As these pieces of information may change over time, the analysis will include data gathered at the time of the last assessment. In this observational cohort, researchers will not control for all of these covariates but will adequately monitor and analyse their influence on the disability outcomes in both groups with or without the exposure factor.

### Blinding and observation bias

Five groups of individuals will be involved in this study: participants, orthopaedists, radiologists, RA, index and reference standard technicians, and statistician. The following actions will be taken to avoid observation bias. The orthopaedists will perform the physical examination blinded to the index or reference standard test results. The radiologists will read the index tests blinded to the reference standard test results, participant identification, physical exam, and laterality of the ankle with the suspected syndesmotic lesion. The radiologists will read the reference standard tests blinded to the participant identification, physical exam results, and index test results. The RA will collect the questionnaires blinded to the index or reference standard tests and the physical exam results. The technician will perform the index tests blinded to the reference standard test or physical exam results. Another technician will perform the reference standard test blinded to the index test or physical exam results. Labelling the variables with nonidentifying terms (such as A and B) will blind the statistician. Researchers will not communicate participants their group assignment.

### Patient and public involvement

Neither the participants nor the public were involved in planning or developing this protocol. Participants will receive information about the knowledge obtained in this study through correspondence or emails. Open lectures in the hospital auditorium and social media communication will disseminate the results to the public.

### Statistical analysis

#### Descriptive analyses

Absolute frequencies and percentages will describe categorical variables. Means, standard deviations, medians, quartiles, minimum, and maximum values will define numerical variables depending on data distribution.

#### Inference analyses of diagnostic accuracy

Sensitivity, specificity, AUC and likelihoods ratios (LR) will be used to compare the diagnostic accuracy of the three strategies. The first strategy will test the diagnostic accuracy of the existing index CTNP alone, the second strategy will examine the add-on index CTSM-EK after CTNP, and the third strategy will assess the add-on index CTSM-FK after CTNP. For statistical analysis, the “either positive” rule will be adopted, whereby the add-on test is positive if either component test is positive. The relative and absolute difference in sensitivity and specificity will compare the performance of the three strategies considering the paired nature of the data. The add-on strategies will reflect superior accuracy if the positive LRs are higher than those of the existing strategy, and the confidence interval (CI) for the LR does not contain 1. The existing strategy will reflect superior accuracy if the negative LR is less than that of the add-on strategies, and the CI for the LR does not incorporate 1.

#### Inconclusive and missing index test data

Inconclusive invalid results of the index tests may occur and will be regarded as uninterpretable when the critical characteristic of the test is corrupted or has questionable validity as a result of the low-quality procedure. The missing index result may occur when a participant is included in the study but does not perform the index test. Missing index results will be treated in the same manner as inconclusive invalid results, and both will be reported separately from the valid results. The underlying causes will be reviewed and discussed, considering the participant’s disease status, an alternative target condition, or whether they are unrelated to the participant. A test is valid (technically appropriate) but inconclusive when the result is neither positive nor negative. For the continuous index test measurements, “rule in” and “rule out” will define thresholds, considering an interval of valid inconclusive test values. The cross-table will consider the number of participants’ positive and negative results for disease in each category, accommodating new groups in extra rows. Paired histograms, dot plots, or cumulative distribution graphs will show the distribution of the test results stratified by disease status. To analyse the valid inconclusive results, multi-level or stratum-specific LR will summarize the performance of the index tests on a quantitative test scale, thus reducing spectrum bias susceptibility.

#### Missing and inconclusive reference standard test data

In this study, a small, incidental, random amount of missing and inconclusive invalid reference standard test data unrelated to the participant characteristics or index test results may occur. The percentage of missing or invalid data will be reported separately from the valid results, including the distribution of participants’ characteristics and index test results in those with or without a reference standard test. A sensitivity analysis will measure the impact of these missing or invalid data. Confirming this scenario, the researchers will perform a complete case analysis, including participants with index and reference standard tests and excluding participants with missing or invalid reference standard tests. If a complete case analysis is found to yield biased accuracy results, an analytical approach will use the available data to reconstruct the missing or invalid data by inverse probability weighting or multiple imputations.

#### Analysis of variability

The subgroup analysis of participants will assess the sources of variability in the index test accuracy. The degree of the severity of the ankle sprain found in the reference standard test will prespecify three subgroups based on the number of syndesmotic ligaments damaged. Isolated lesions of the anterior tibiofibular ligament will identify a mild sprain. Injuries involving the anterior tibiofibular and interosseous ligaments will define a moderate sprain. Lesions involving the anterior tibiofibular, interosseous, and posterior tibiofibular ligaments will indicate a severe sprain. The index test performance is expected to be more accurate with the higher degrees of sprain than with the lower degrees of sprain. Pain exacerbation during the stress manoeuvre may be another source of variability. Pain aggravation reported by the participant will prespecify one subgroup and pain non-aggravation the other subgroup. The pain aggravation subgroup may have difficulty performing dorsiflexion, and lower accuracy results compared to those in the pain non-aggravation subgroup may occur. Sensitivity, specificity, AUC, and LR will be used to examine the diagnostic accuracy of the subgroups.

#### Interrater analysis

Two observers will independently read the index tests, and two other observers will independently read the reference tests. The intraclass correlation coefficient will verify the agreement between observers regarding the data extracted from the index test, and the Kappa coefficient will confirm the agreement between observers related to data derived from the reference standard test. A second consensus reading will be used to solve discordant cases.

#### Inference analyses for the FAAM questionnaire

Mixed linear models or generalized linear models in multiple approaches will evaluate the association of the SI findings, participants’ characteristics (age, gender, and BMI) and treatment (weight-bearing restriction, immobilization, physical rehabilitation, and surgery) with the change in the disability (FAAM) at six and 12 months after CT. These models are appropriate for measurements obtained from the same participant at different times. The effect estimates will present the results (change in mean functional disability) with 95% confidence intervals. A two-sided t-test will compare the changes in the FAAM score between the groups.

#### Sample size calculation

The sample size calculation will consider the comparison of the primary objective (diagnostic accuracy) and the secondary objective (change in the FAAM disability score). The most significant sample needed to meet the primary or secondary objectives will define the study sample size.

A previous study found an AUC performance of 0.56 relative to the CTNP.^12^ Assuming as a null hypothesis that the existing CTNP test has an AUC of 0.56, this study proposes as an alternative hypothesis a new CTSM test with superior accuracy, which is estimated as an AUC of 0.80. The full sample size required to detect a difference between these two is estimated as 39, with a 1:2 allotment ratio between the groups (13 and 26 cases per group, respectively).

A previous study found that the effect size of the change score paired data for the FAAM Sports subscale was 15, with a standard deviation of change of 28 in one group and 23 in the other group.^22^ Using a two-sample paired t-test to compare the changes in the Sports subscale, the required full sample size is 111, with a 1:2 allocation ratio between the groups (37 and 74 cases per group, respectively). Based on the same study^22^ and applying the same methodology, the required full sample size for the ADL subscale is 60 participants.

The sample of 111 individuals required for the Sports subscale defined the study sample size because contemplates the sample calculations of the ADL subscale and the AUC. Accounting for a possible loss of 20% during follow-up, the investigators intend to include a total of 133 participants in the study.

Once the study has reached a follow-up of 67 participants, the sample size needed to compare the groups will be recalculated based on both the standard deviation and the proportion of syndesmotic lesions observed in this study.

#### Software and thresholds

Program R (R Core Team, 2017), version 3.4.1, will perform the analyses, with 80% power to test group differences at a significance level of 5% and 95% confidence intervals.

## ETHICS AND DISSEMINATION

The Internal Review Board approved this study, which will be conducted according to the requirements of the Research Ethics Committee (62100016.5.0000.0071) and based on the recommendations established by the Declaration of Helsinki (2013) and Document Guidelines for the Americas. Participation is free and voluntary, and all recruited individuals will receive a verbal explanation of the purpose of the study. This study will enrol participants after they have signed the informed consent form, including confidentiality terms. To disseminate the knowledge, researchers will present the results at national and international conferences and will publish the results in peer-reviewed scientific journals and social media.

## DISCUSSION

Although there are several imaging options for the diagnosis of syndesmotic injury, a fundamental question guiding treatment remains unanswered. Currently, imaging tests readily diagnose the severe SI but have difficulty in diagnosing mild and moderate cases. Presumably, the correct treatment is not being applied to a significant proportion of individuals. Undiagnosed and untreated mild and moderate SI are considered the primary source of unfavourable outcomes. If this study confirms CTSM as an accurate method of diagnosing SI, a new approach to investigate challenging cases may be opened, and perhaps more individuals will benefit from correct treatment, reducing the burden of unfavourable outcomes. An algorithm combining clinical suspicion, MRI findings, and CTSM protocol may be the correct and precise way to diagnose SI.

### Strengths

To the best of our knowledge, this study is the first to test the CTSM examination accuracy for SI and to test the feasibility of stress manoeuvres, which will be evaluated based on the pain perception reported by participants in addition to the objective parameters measured by the performing technician. This study is also the first to test SI, diagnosed with CTSM, as a prognostic factor for disability outcomes. This assessment is not confined only to exposure (with or without SI). Participants’ demographics and treatment characteristics will also be investigated for their influence on the outcomes.

### Limitations

This study uses MRI as the reference standard test, which, although not perfect, has a high estimated accuracy compared to the gold-standard arthroscopy.^1^ An inherent degree of imprecision related to the severity of the ankle sprain and pain exacerbation may occur when the participants themselves control stress manoeuvres; therefore, some variability is unavoidable. The researchers will monitor and investigate the influence of this source of variability in the results. A loss-to-follow-up bias may occur because the unexposed group is expected to have more pronounced symptom relief, which may prevent individuals from participating until the end of the follow-up. The researchers will use a questionnaire that participants can answer at home, by phone, or online and will reinforce that the study is essential, regardless of an improvement in or aggravation of symptoms, to maintain the participants’ interest and minimize this effect.

## Data Availability

The actual publication is a study protocol with no data to present. In a forthcoming paper with the results, researchers intend to share data.

## Acknowledgments

The authors gratefully acknowledge the contribution of Gilberto Szarf for providing all the support to turn an idea into a real project; Leonardo Kamibeppu and Fabio Augusto for all help with biomedical activities; Elivane da Silva Victor and Ana Carolina Cintra Nunes Mafra for the valuable contribution in the statistical plane; Rosemeire Pereira Bezerra and Giovanna de Souza Mendes for all help with administrative issues and organization of the questionnaires, and Maria Angela M. Barreiros for production of the flowchart.

## Contributors

JCR conceived the study, participated in its conception, design, coordination, and drafted the manuscript. ALGS and MPP participated in the conception and design of the project and writing the paper. RAM, JFMA, DCSB and LAR participated in the design and coordination of the study and contributed to the critical revision of the article. AAC, MKD, ML and MF participated in the design and critical review of the manuscript. All authors read and approved the final version of the paper.

## Participant consent

All participants will receive a verbal explanation about the purpose of this study, and the decision to participate is free and voluntary. The participants’ enrollment will only occur after the written informed consent, including confidentiality terms, has been signed.

## Ethics approval

The Internal Review Board and the Research Ethics Committee (register number 5291417.0.0000.0071) approved this study that will be conducted based on the recommendations established by the Declaration of Helsinki (2013) and Document Guidelines for the Americas.

